# Health impact of routine measles vaccination and supplementary immunisation activities in 14 high burden countries: a DynaMICE modelling study

**DOI:** 10.1101/2022.07.11.22277494

**Authors:** Megan Auzenbergs, Han Fu, Kaja Abbas, Simon R Procter, Felicity Cutts, Mark Jit

**Affiliations:** Department of Infectious Disease Epidemiology, London School of Hygiene & Tropical Medicine, London, UK; Public Health Foundation of India, New Delhi, India; International Vaccine Institute, Seoul, South Korea; School of Public Health, University of Hong Kong, Hong Kong, SAR, China

## Abstract

**Background:** WHO recommends ≥95% population coverage with two doses of measles-containing vaccine (MCV). Most countries used routine services to offer MCV1 and later, MCV2. Many countries conducted supplementary immunisation activities (SIAs), offering vaccination to all persons in a given age range irrespective of prior vaccination history. We estimated the relative impact of each dose and delivery route in 14 high burden countries.

**Methods:** We used an age-structured dynamic model (DynaMICE), to estimate the health impact of different vaccination strategies on measles susceptibility and burden over 2000-2020. We estimated their incremental impact using averted cases and deaths and their efficiency using number needed to vaccinate (NNV) to avert an additional measles case.

**Findings:** Compared to no vaccination, MCV1 implementation averted 823 million cases and 9.5 million deaths, with a median NNV of 1.41. Adding routine MCV2 to MCV1 further averted 108 million cases and 0.4 million deaths, while adding SIAs to MCV1 led to 249 million averted cases and 4 million deaths. Despite a larger incremental impact, adding SIAs to MCV1 showed reduced efficiency compared to adding routine MCV2, with median NNVs of 6.15 and 5.41, respectively.

**Interpretation:** Vaccination strategies including non-selective SIAs reach a greater proportion of unvaccinated children and reduce burden more than MCV2 alone, but efficiency is somewhat lower because of the wide age groups included in SIAs. This analysis provides insight to improve health impact and efficiency of measles vaccination.

**Funding:** Gavi, the Vaccine Alliance, and the Bill & Melinda Gates Foundation (OPP1157270)

## Introduction

Between 2000–2020, measles deaths were estimated to have decreased by 94% globally [1], achieved mostly through routine immunisation (RI) and supplementary immunisation activities (SIAs) with measles-containing vaccines (MCV) [2-5]. MCV1 is defined by the World Health Organization (WHO) as the first routine dose of MCV given during the first year of life (recommended at 9 or 12 months of age) while MCV2 is defined as the second routine dose of MCV (recommended at 15–18 months of age). SIAs refer to vaccination campaigns that deliver vaccine doses using strategies going beyond routine services and have usually been non-selective, that is offering vaccine irrespective of past vaccination history. Throughout this paper, the term SIA indicates non-selective SIAs.

Since the introduction of measles vaccination in low- and middle-income countries (LMICs), recommendations around measles vaccination strategies have been revised. Historically, LMICs relied on MCV1 with SIAs to interrupt transmission and reach unvaccinated children. In 2009, WHO recommended introducing MCV2 once a country reached 80% MCV1 coverage, retaining an emphasis on aiming for high coverage with MCV1 as soon as possible after a child loses maternal antibodies. In 2017, this policy was revised to recommend that countries include MCV2 in RI schedules regardless of MCV1 coverage alongside operational support to strengthen RI infrastructure when incorporating MCV2. In part due to concerns about the sustainability of funding for nationwide non-selective SIAs and their potential to disrupt routine services [6, 7], WHO has proposed that eventually such SIAs can be phased out once countries achieve over 95% coverage of both routine doses [8]. As of 2021, 179, or 92%, of countries have incorporated MCV2 in their routine programmes, although no WHO region has collectively achieved over 95% MCV2 coverage and disparate levels of coverage persist across Sub-Saharan Africa, Latin America & Caribbean, and East Asia & Pacific regions [9, 10].

Implementation of SIAs over time has been motivated by different goals and needs. For example, SIAs have been used to increase population immunity in countries with low MCV1 or MCV2 coverage. In such settings, SIAs have been cited as a highly effective and equitable strategy for protecting hard-to-reach communities with children who would otherwise be missed by RI [11, 12], although the relative reach of SIAs versus RI varies between and within countries [13]. Importantly, SIAs were a major component of the measles elimination strategy in the region of the Americas, with high routine MCV1 coverage and occasional follow-up SIAs sustaining elimination for many years [14]. To prevent measles transmission and subsequent outbreaks, a commonly used (rule-of-thumb) criterion is that a follow-up SIA should be conducted before the number of susceptible children under 5 years of age approaches the size of one birth cohort [15, 16]. Historically, this rule-of-thumb has been influential in informing the timing of SIAs so the accumulation of susceptibles remains below the size of one birth cohort and measles transmission can be interrupted and elimination achieved [8]. In practice, even if countries recognise that a follow-up SIA is due and correctly identify the age groups with the highest prevalence of susceptibility, delays in obtaining funding or competing priorities may lead to delayed implementation of an SIA or a narrower than ideal age range, which reduces its impact [16].

In 2012, the World Health Assembly endorsed the Global Vaccine Action Plan, which included a commitment to achieving measles elimination in five of the six WHO regions by 2020. From 2000–2010, estimated global MCV1 coverage increased from 72% to 84%, but has stagnated since, although estimated routine MCV2 coverage has increased from 18% in 2000 to 70% in 2020 [17]. In 2020 during the COVID-19 pandemic, more than 27 million infants worldwide were estimated to have missed their first MCV dose [18, 19]. As priorities shifted and resources were diverted to support the COVID-19 response, measles surveillance systems deteriorated, and disease monitoring and testing was weakened [20, 21]. Therefore, as health systems rebuild following the pandemic, it is important to understand historical vaccination policies and shortcomings to make informed decisions about future vaccination programmes.

This retrospective analysis of measles vaccination policies from 2000–2020 uses the Dynamic Measles Immunization Calculation Engine (DynaMICE), a population-based dynamic model of measles transmission, to better understand the impact of different vaccination strategies that have been used in 14 high burden countries.

## Methods

### Data sources

Reported measles cases collected through the WHO/UNICEF Joint Reporting Form on Immunization and estimated measles incidence data from the Institute for Health Metrics and Evaluation (IHME) were used to obtain separate rankings of countries by measles incidence from 2010–2019[22, 23]. We included the top 10 countries from each data source (appendix table S1), which resulted in a total of 14 unique countries included in the analysis: India, Nigeria, Indonesia, Ethiopia, China, Philippines, Uganda, Democratic Republic of the Congo (DRC), Pakistan, Angola, Madagascar, Ukraine, Malawi, Somalia—contributing to 35% of the global birth cohort from 2012–2020.

Country-specific routine MCV1 and MCV2 coverage data from 1980–2020 were obtained from the WHO and UNICEF Estimates of National Immunization Coverage (WUENIC) database for all modelled countries [24] and SIA data were obtained from the WHO summary of measles and rubella SIAs (figure 1) [25]. We extracted the year and month of implementation, targeted age group, and the number of doses given during each SIA. Many countries, such as DRC and India, conducted national SIAs in a phased manner over 2-3 years, while others undertook subnational SIAs in specific high-risk areas. It is not always possible to know whether the entire country was covered after phased or subnational SIAs, hence in figure 1, for each year, we calculated country-level coverage by comparing reported SIA doses to the entire national population in the target age range.

**Figure 1.**
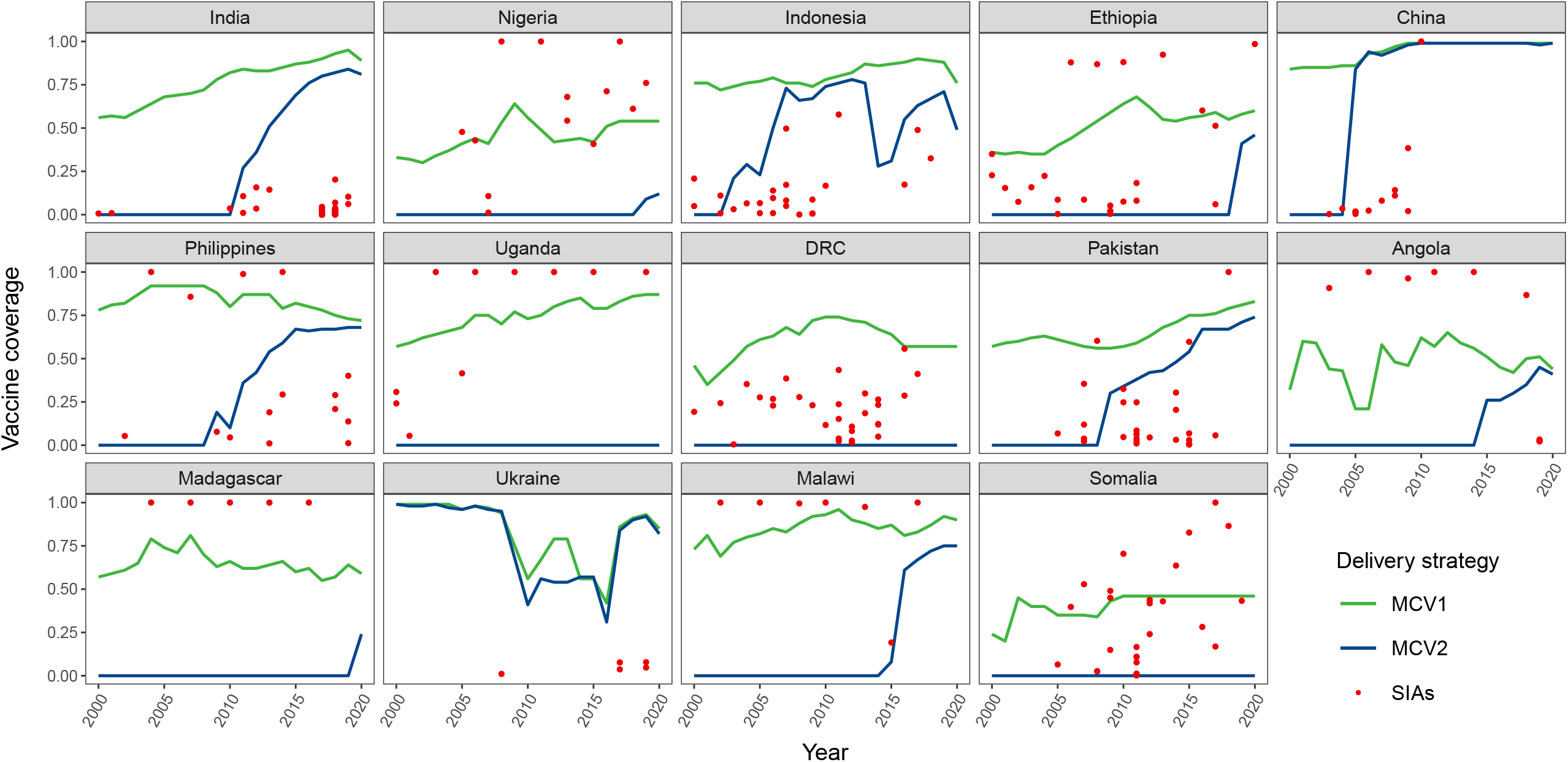
Immunisation coverage for MCV1, MCV2, and SIAs over 2000–2020. The calculation of SIA coverage was based on reported numbers of doses administered and national populations in the SIA target age group. DRC: Democratic Republic of the Congo. MCV1: the first routine dose of measles-containing vaccine. MCV2: the second routine dose of measles-containing vaccine. SIA: supplementary immunisation activity.

### DynaMICE Model

DynaMICE is an age-structured compartmental model of measles transmission that considers time-varying states of disease (maternally immune, susceptible, infectious, recovered) and vaccination (0, 1, 2, ≥3 doses). The model has been used previously for estimating the impact of measles vaccination [2, 7] and a detailed description of the model structure, parameters, and equations has been published [26]. Utilising the DynaMICE model, we modelled country RI programmes based on historical WUENIC coverage estimates and nationally recommended schedules for MCV1 and MCV2 (appendix table S1) [23]. We assumed SIA doses are more likely to reach previously vaccinated children, by distributing doses randomly among the target population except for a proportion who are never reached by measles vaccination programmes (derived from the estimate for children missing all essential childhood vaccination, comprising an average of 7.7% of 1-year old children in LMICs over the period 2010–20) [27] (figure S2). In the model, MCV1 efficacy increases linearly with age, from 74% at 6 months old up to a maximum of 98% at 23 months, the level of protection provided by two-dose vaccination [28, 29]. The basic reproduction number, R0, was 15.9, based on a summary estimate taken from endemic settings [30]. Country and age-dependent social contact matrices [31] were used to inform the country-specific patterns of measles transmission. To capture the epidemic trend since the wide implementation of MCV, the simulation began in 1980. The code can be accessed at: https://github.com/hfu915/dynamice_ph.

### Measles vaccination scenarios and impact estimates

Using the DynaMICE model, we assessed (undiscounted) measles cases, deaths, and disability-adjusted life years (DALYs) over 2000–2020 across varying vaccination scenarios: (i) no vaccination, (ii) MCV1, (iii) MCV1 + MCV2, (iv) MCV1 + SIAs, and (v) MCV1 + MCV2 + SIAs. We estimated deaths by multiplying the model estimates of cases with age-, year- and country-specific case-fatality ratios [32]. DALYs include years of life lost due to measles deaths, defined as the country-specific remaining life expectancy at age of death, and years of disability due to acute morbidity based on a disability weight of 0.051 for moderate severity infections [22] over an illness period of 14 days. We calculated the annual incidence per 1 million population and compared the under-five susceptible population to the birth cohort, defined as the mid-year population aged between 0 and 1 year old, to understand the potential of different delivery strategies to reduce transmission and outbreaks. To estimate the incremental impact of historical measles vaccine strategies, each scenario was compared to a counterfactual scenario, representative of a historical policy decision for measles vaccination. MCV1 was compared to the alternative of no vaccination (scenario ii vs i) while MCV1 + MCV2 (scenario iii vs ii) and MCV1 + SIAs (scenario iv vs ii) were compared to MCV1 alone. MCV1 + MCV2 + SIAs was compared to the counterfactual scenario MCV1 + SIAs (scenario v vs iv), and separately, compared to MCV1 + MCV2 (scenario v vs iii). For each pair of comparison, we estimated the ‘health impact’ of an additional delivery strategy based on the cumulative vaccine-averted cases, deaths, and DALYs over 2000–2020, and the ‘efficiency’ of adding a delivery strategy through the number needed to vaccinate (NNV) to prevent a measles case. Since DRC, Uganda, and Somalia had not introduced MCV2 by 2020, strategies including MCV2 were not assessed for these 3 countries.

### Sensitivity analysis

In this alternative scenario, we modelled the vaccine impact if MCV2 had been introduced early in 2000. For each year from 2000 to 2020, we assumed that the alternative MCV2 coverage was either 10% lower than the country’s MCV1 coverage or equal to the country’s MCV2 coverage in that year, whichever was larger (dashed lines in appendix figure S1). In addition, we modelled two alternative assumptions about the likelihood of receipt of an SIA dose according to past vaccination history (appendix figure S2). In this analysis, we specifically describe ‘zero-dose’ population as those receive no MCV doses. One assumption is that SIA doses preferentially reached already-vaccinated children, and any remaining doses after all already-vaccinated children are reached are then given to zero-dose children, while the other assumes a strategy that reaches zero-dose children first, and the remaining doses are given to already-vaccinated children. While these two distribution assumptions are hypothetical, these analyses strengthen our understanding on the potential ranges of incremental health impact and efficiency that SIAs can provide when added to MCV1 only strategy.

## Results

Figure 2 illustrates estimated measles incidence rates per million population from 2000-2020 across different vaccination delivery scenarios, based on the country-specific WUENIC coverage estimates and SIAs reported to WHO. Compared to the no vaccination scenario, all 14 high-burden countries presented a substantial decline in incidence rates in scenarios with MCV1 only. In the scenario where MCV1 and MCV2 were used without SIAs, the annual burden of measles declined slowly over time, and endemic transmission continued. With MCV1 and SIAs, there was a more rapid decline in measles burden, but large-scale outbreaks were predicted. Overall, the greatest absolute burden reduction attributable to MCV1, MCV2, and SIAs in comparison to no vaccination over 2000–2020 was in India, China, and Nigeria respectively (appendix figure S3), which are countries with the highest IHME measles incidence estimates and largest population sizes.

**Figure 2.**
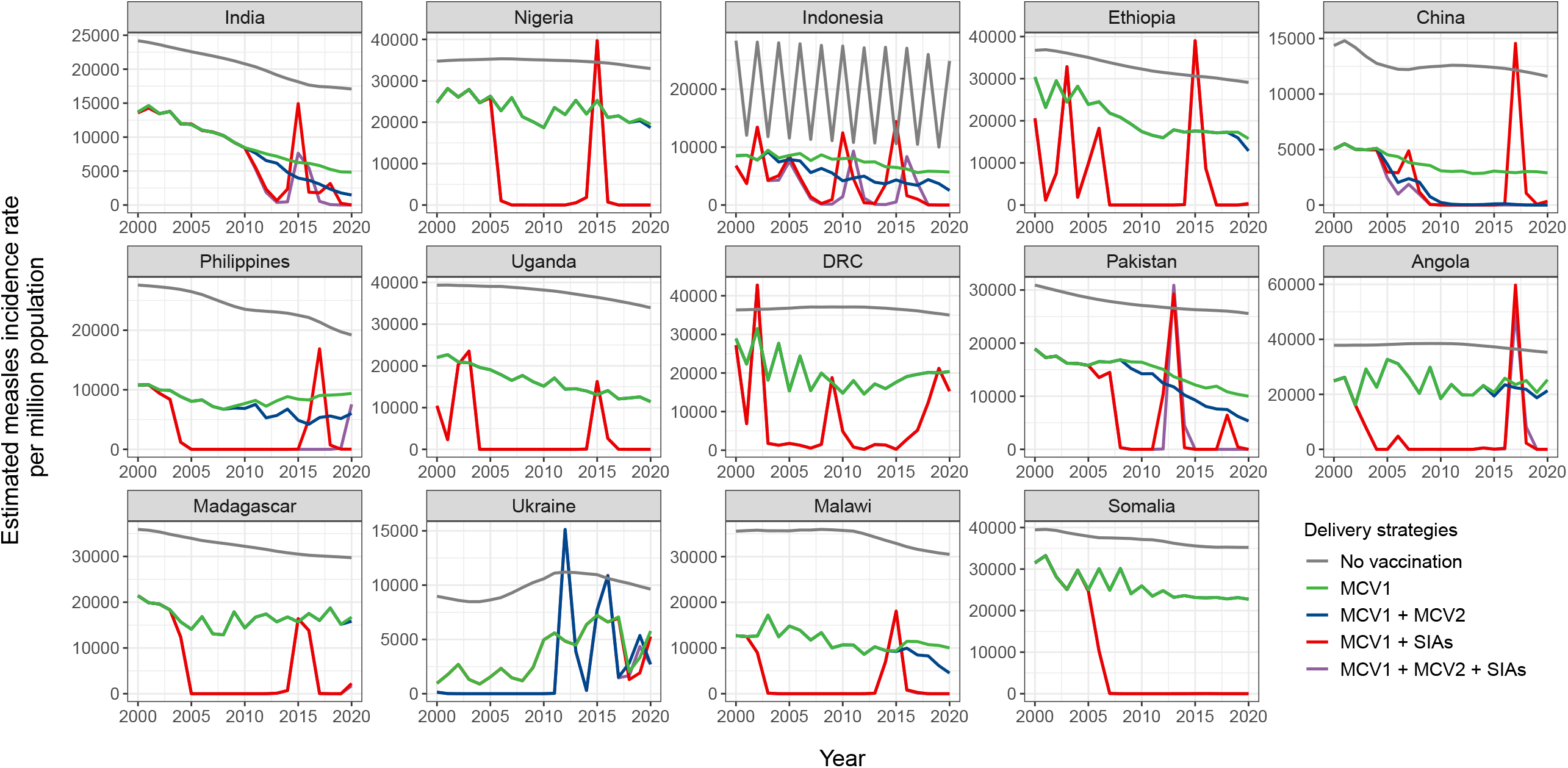
Estimated annual measles incidence rate per million population across different vaccination delivery strategies over 2000–2020. In each country, temporal trends of measles incidence rates vary by different vaccination delivery strategies (coloured lines). The measles burden decreases with adding vaccination delivery strategies. For countries that have not yet introduced MCV2 (Uganda, DRC, and Somalia), there are overlapping trends for incidence rates under the strategies of MCV1 and MCV2 (blue lines) and MCV1 only (green lines). DRC: Democratic Republic of the Congo. MCV1: the first routine dose of measles-containing vaccine. MCV2: the second routine dose of measles-containing vaccine. SIA: supplementary immunisation activity.

The estimated total number of susceptible children under 5 years of age shows varying trends by vaccination delivery strategy (figure 3). Routine immunisation with MCV1 and MCV2 reduced measles susceptibility compared to the counterfactual scenario with no vaccination, but the numbers of susceptible children remained greater than one birth cohort in 11 of the analysed countries over 2000–2020. China was an exception to this trend, where high MCV1 and MCV2 coverage has successfully kept the susceptible population under the threshold of one birth cohort since 2007. Despite several rebounds of the susceptible population over the study years, MCV1 and SIAs had more potential in reducing susceptibles compared to MCV1 and MCV2; however, we estimated that in four countries, MCV1 and SIAs would not have reduced susceptibles to below the birth cohort in any year (appendix table S4). Overall, measles vaccination strategies as reported by these countries were estimated to have reduced the number of susceptible children below the birth cohort in a median of 26% (25^th^ and 75^th^ percentiles: 14%-37%) of years between 2000-2020.

**Figure 3:**
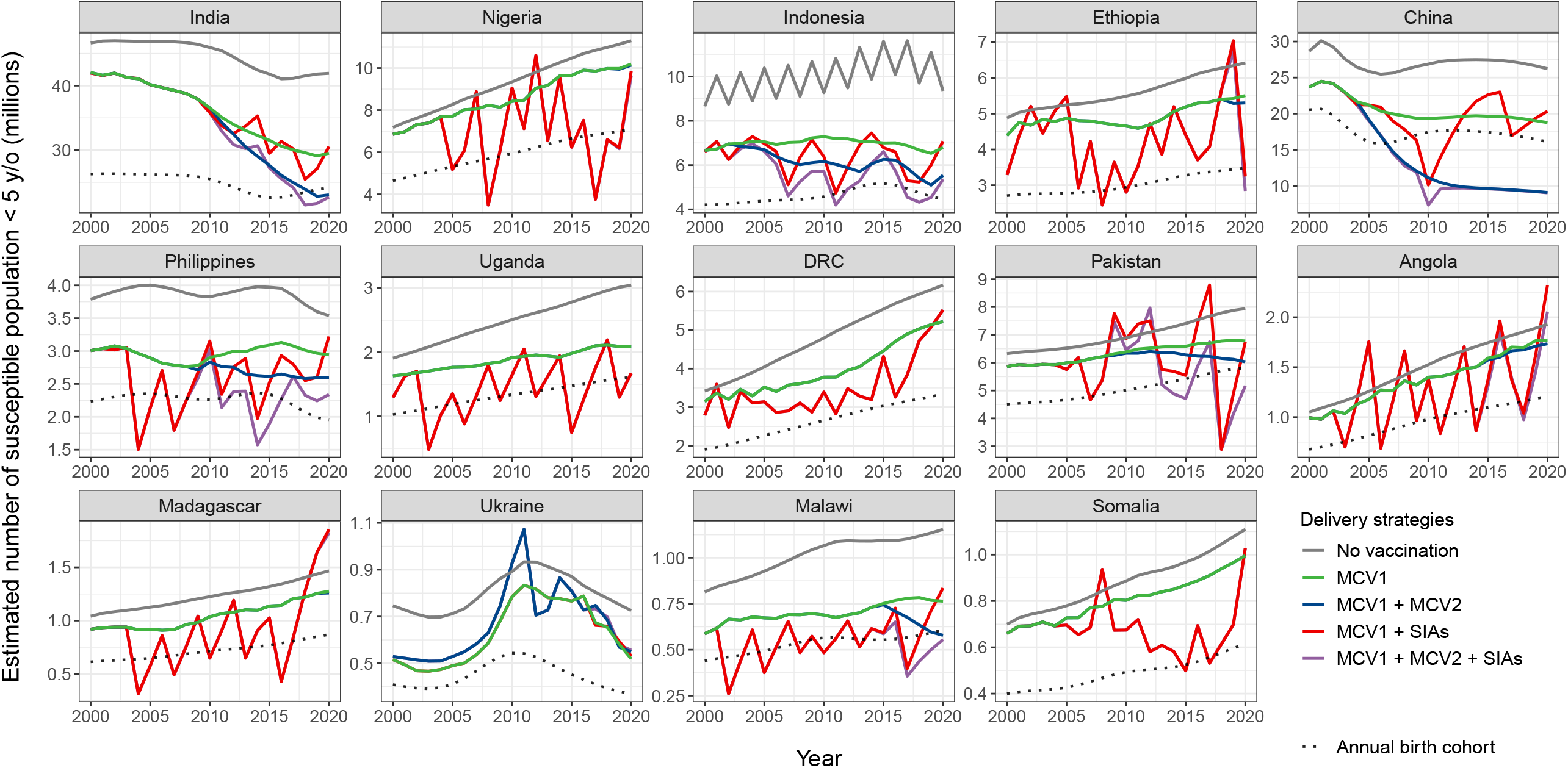
Susceptible population under 5 years old by vaccination delivery strategies over 2000–2020. Estimated total numbers of susceptible persons who are under 5 years old under different vaccination delivery strategies (colour solid lines) are compared with the size of birth cohort (dotted lines). DRC: Democratic Republic of the Congo. MCV1: the first routine dose of measles-containing vaccine. MCV2: the second routine dose of measles-containing vaccine. SIA: supplementary immunisation activity.

Compared to no vaccination, we estimated that MCV1 alone averted 824 million cases and 9.6 million deaths over 2000–2020 in the 14 high-burden countries (table 1 and appendix table S2). The marginal benefit of SIAs done in these countries was a further 249 million cases and 4.07 million deaths averted compared to MCV1 alone. MCV2, as used by these countries, was estimated to have averted 108 million cases and 0.40 million deaths in comparison to MCV1 alone. SIAs demonstrated higher ‘impact’ on the aspect of burden reduction than MCV2 when added to MCV1, as seen in the greater number of averted cases in all countries except China and Ukraine. SIAs also had a greater effect on deaths and DALYs (appendix figure S3). While China had the greatest burden reduction attributable to MCV1 and MCV2 in comparison to MCV1 alone, Nigeria had the greatest burden reduction attributable to MCV1 and SIAs. In addition, both MCV2 and SIAs as used were predicted to have incrementally averted 303 million cases and 4.3 million deaths compared to the scenario with MCV1 alone. We did not observe a synergistic effect of MCV2 and SIAs on burden reduction. The relative impact of different vaccination strategies on averted DALYs followed the trends seen in averted cases and deaths (appendix table S3).

**Table 1.** Averted cases (thousands) across different vaccination delivery strategies as reported by each country from 2000–2020. Five pairs of scenarios were compared to two comparator scenarios to assess the health impact of additional vaccination delivery strategies as reported by each country. MCV2 impact begins in the year when WUENIC first reports MCV2 coverage (table S1). Column one represents ‘total impact’ of MCV1, MCV2 and SIAs in comparison to no vaccination, however, note that this does not depict an actual historic policy implementation for vaccination strategies. Countries are presented in an order according to their introduction year of MCV2 and magnitude of measles burden (table S1). Sums of the averted cases in the 14 countries are presented in the last row of the table. Note that entries with no value listed correspond to options involving MCV2 in the 3 countries (Uganda, DRC and Somalia) that have not yet introduced MCV2, and therefore cases averted cannot be estimated. Since MCV2 had no contribution to burden reduction in these 3 countries, the NNV values are the same between the ‘MCV1 + MCV2 + SIAs’ scenario and ‘MCV1 + SIAs’ scenario when the comparator is the ‘MCV1 alone’ scenario. DRC: Democratic Republic of the Congo. MCV1: the first routine dose of measles-containing vaccine. MCV2: the second routine dose of measles-containing vaccine. SIA: supplementary immunisation activity. WUENIC: WHO and UNICEF Estimates of National Immunization Coverage.

| Country by MCV2 introduction year | Comparator: no vaccination |  | Comparator: MCV1 alone |  |  |
| --- | --- | --- | --- | --- | --- |
|  | MCV1 + MCV2 + SIAs | MCV1 alone | MCV1 + SIAs | MCV1 + MCV2 | MCV1 + MCV2 + SIAs |
| **MCV2 < 2017** |  |  |  |  |  |
| India | 349879 | 295563 | 39541 | 28182 | 54316 |
| Indonesia | 82723 | 61350 | 17251 | 9902 | 21373 |
| China | 321750 | 257140 | 34020 | 57289 | 64610 |
| Philippines | 42782 | 29887 | 11356 | 3164 | 12895 |
| Pakistan | 76702 | 49163 | 25279 | 7070 | 27539 |
| Angola | 15586 | 6780 | 8689 | 433 | 8805 |
| Ukraine | 7321 | 6090 | 125 | 1132 | 1232 |
| Malawi | 9725 | 7003 | 2715 | 313 | 2722 |
| **MCV2 > 2017** |  |  |  |  |  |
| Nigeria | 88243 | 39343 | 48900 | 251 | 48900 |
| Ethiopia | 48865 | 23045 | 25794 | 484 | 25820 |
| Madagascar | 12044 | 6935 | 5095 | 25 | 5109 |
| **No MCV2** |  |  |  |  |  |
| Uganda | 23867 | 15024 | 8843 | 0 | 8843 |
| DRC | 39662 | 23429 | 16233 | 0 | 16233 |
| Somalia | 7632 | 2931 | 4702 | 0 | 4702 |
| **Total** | **1126781** | **823682** | **248544** | **108245** | **303099** |

Table 2 shows NNV estimates for each scenario compared to different counterfactual scenarios, showing the number of additional doses that need to be given to prevent a measles case. Compared to no vaccination, the NNVs for MCV1 ranged between 1.27 and 1.46. In comparison to MCV1 alone, the way SIAs had been used had a median NNV of 6.15, which was greater than adding MCV2 in 7 of the 11 countries that have introduced MCV2 (median NNV = 5.41). The opposite trend of NNV was observed in Nigeria, Ethiopia, Angola, and Madagascar, where frequent SIAs took place and MCV2 was only introduced recently (appendix table S1). Furthermore, adding SIAs when both MCV1 and MCV2 were used led to a median NNV of 6.49, while adding MCV2 when both MCV1 and SIAs were used resulted in a median NNV of 14.3. There is diminishing returns in efficiency for including an additional vaccination delivery strategy when multiple strategies have been already in use.

**Table 2.**
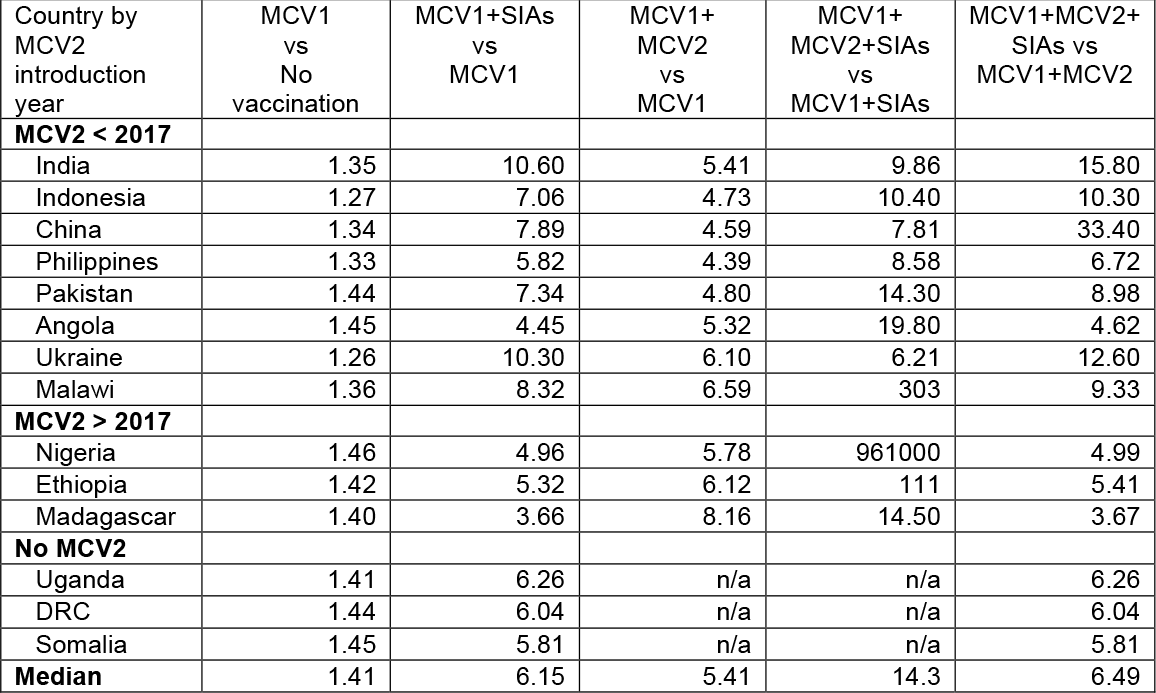
Number needed to vaccinate (NNV) to avert a measles case across different vaccination delivery strategies over 2000–2020. Number needed to vaccinate (NNV) is defined as the ratio of incremental averted cases to incremental doses in a vaccine delivery strategy compared to its comparator. The median NNVs among countries with applicable values for the 5 comparison pairs are presented in the last row of the table. Note that entries with no NNV value listed correspond to options involving MCV2 in the 3 countries (Uganda, DRC and Somalia) that have not yet introduced MCV2, and therefore NNV cannot be estimated (denoted as ‘n/a’). Since MCV2 had no contribution to burden reduction in these 3 countries, the NNV values are the same between the ‘MCV1 + MCV2 + SIAs vs MCV1 + MCV2’ scenario and ‘MCV1 + SIAs vs MCV1’ scenario. For the ‘MCV1 + MCV2 + SIAs vs MCV1 + MCV2’ scenario, the NNV in Nigeria is exceptionally large due to a small number of averted cases from recent MCV2 introduction in 2019. DRC: Democratic Republic of the Congo. MCV1: the first routine dose of measles-containing vaccine. MCV2: the second routine dose of measles-containing vaccine. NNV: number needed to vaccinate to avert a case. SIA: supplementary immunisation activity.

In figure 4, we estimated averted cases and NNVs under alternative assumptions for MCV2 introduction and SIA dose distribution. First, instead of following the historical introduction schedule, we explored the scenario if MCV2 were introduced earlier in 2000 with a coverage trend being 10% lower than MCV1 (appendix figure S1). Compared to MCV1 alone, early introduction of MCV2 would have averted more cases than the historical introduction, resulting in a further reduction of 97 million measles cases across study countries. In terms of efficiency, only slight improvement was seen from early MCV2 introduction, with a median NNV dropping from 5.41 to 5.09. Next, we explored the SIA dose distribution approaches that allow doses to be delivered firstly to zero-dose or already-vaccinated children, while in the main analysis, 7.7% of population are assumed to be never reached by measles vaccination and SIA doses are distributed randomly among the remainder (figure S2). We demonstrated that the distribution of SIA doses between zero-dose and already-vaccinated populations had a strong effect on the incremental impact and efficiency of vaccination. When MCV1 was already in use, successfully directing SIA doses first to zero-dose children and the remainder to already-vaccinated children could prevent more cases than early introduction of MCV2 in all countries, except China. As a strategy with substantial impact on case reduction, prioritising the zero-dose population for SIA doses improved efficiency (median NNV = 4.55 compared to 6.15 in the main analysis), for countries with a gap in routine immunisation coverage, such as Nigeria and DRC. Conversely, when SIA doses first reached already-vaccinated children, the median NNV increased to 8.34 and substantially reduced the number of averted cases compared to the main analysis.

**Figure 4.**
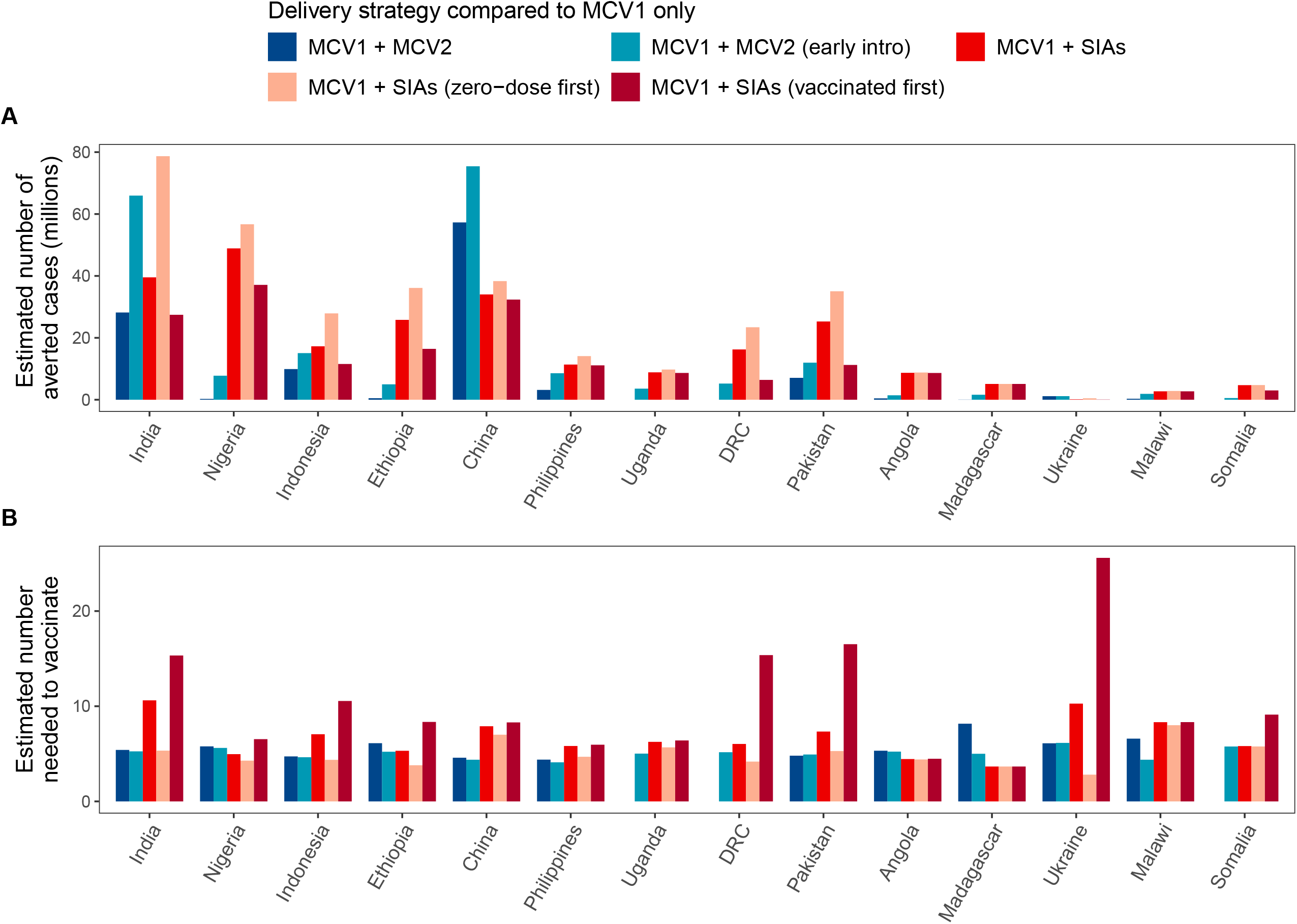
Averted cases (A) and number needed to vaccinate to avert a measles case (B) under alternative assumptions for early MCV2 introduction and different SIA dose distribution. In the sensitivity analysis, we modelled the incremental impact and efficiency of vaccination under the alternative scenarios of MCV2 introduction and SIA distribution. The incremental impacts of all the scenarios are compared to the scenario where MCV1 was already in use. In the main analysis, MCV2 was introduced based on its historical WUENIC coverage (dark blue) and SIAs were distributed with an assumption that 7.7% of children were never reached by vaccination (red). The alternative MCV2 scenario indicates early introduction of MCV2 in 2000 with coverage inputs from figure S1 (light blue). Three countries (Uganda, DRC, and Somalia) that have not yet introduced MCV2 have missing estimates for the original scenario with MCV1 and MCV2 in the figure. Two alternative scenarios for SIA distribution were evaluated, including prioritisation to zero-dose children (pink) and reaching firstly vaccinated children (dark red), respectively. DRC: Democratic Republic of the Congo. MCV1: the first routine dose of measles-containing vaccine. MCV2: the second routine dose of measles-containing vaccine. SIA: supplementary immunisation activity.

## Discussion

Using 20 years of MCV coverage data, we investigated the relative impact and efficiency of different MCV strategies in 14 high measles burden countries that included a wide range of socio-economic, demographic and routine immunisation settings. We describe health impact in terms of cases, deaths, and DALYs averted, and the proportion of years during which vaccination scenarios kept the susceptible population below one birth cohort, while efficiency is described in terms of NNV to avert a measles case.

The use of MCV1 resulted in the highest relative health impact of any dose, and the best efficiency in reducing measles burden. The vaccination scenario using MCV1 and SIAs together can more effectively keep the size of the susceptible population below the size of one birth cohort and had a bigger effect on predicted incidence in comparison to using MCV1 and MCV2 together. Furthermore, SIAs averted a greater proportion of measles burden and reduced the size of the susceptible population more than MCV2, whereas efficiency of SIAs as assessed by NNV to avert a measles case was lower than that for MCV2. However, there was marked variation between countries in the relative efficiency of each incremental strategy. The strategies used between 2000–2020 in these countries greatly reduced measles burden compared to a no-vaccination scenario but, other than in China, were not predicted to prevent large outbreaks. This is consistent with other analyses in LMICs [33].

The relatively high impact but reduced efficiency of SIAs could also be interpreted from the viewpoint of dose delivery ― while SIAs could be delivered to more people, many doses were predicted to reach previously vaccinated children (appendix figure S4). Even though prioritisation of zero-dose children for measles vaccination during SIAs may not always be the most *efficient* strategy as it is operationally challenging and costly, successfully reaching zero-dose children first results in an *effective* strategy to avert measles burden.

Compared to SIAs, MCV2 had relatively less impact in countries that had not yet met or exceeded 90% routine coverage of MCV1, even under the assumption of early MCV2 introduction. Accompanied by better reach of zero-dose children by SIAs, early introduction of MCV2 could have substantially reduced incidence over time. In countries that implemented routine MCV2 early on and maintained high levels of coverage, such as China and India, there was little difference in estimated incidence rate between historical scenarios and “optimal” assumptions for MCV delivery that MCV2 was introduced in 2000 and SIA doses were given first to zero-dose children (appendix figure S5). This finding suggests that in the future SIAs may be needed less often if high coverage of MCV2 can be successfully attained and maintained and aligns with the recommendations outlined in the WHO position paper on measles vaccines [8].

The Measles and Rubella Strategic Framework 2021–2030 emphasises shifting from a ‘one size fits all approach’ to focus on effective local approaches for vaccinating hard-to-reach populations with MCV [34]. To address the strategic framework’s concern for unvaccinated populations, the results from this research need to be considered when planning vaccination programmes. For example, if SIA efficiency is low in a particular setting, as shown by a high predicted NNV to avert a case, but SIAs consistently result in a greater burden reduction than MCV2, then investing in mechanisms to improve efficiency may be worthwhile.

There are limitations in capturing the relative health impact and efficiency of different vaccination strategies in this study. First, SIA doses were assumed to be randomly delivered to their target population, except for a fixed proportion of children who were assumed never reached by childhood immunisation programme [27]. The extent to which in practice, doses are correlated with, or independent of, previous vaccination status is unknown because only a minority of countries report high-quality post-campaign-coverage surveys to WHO and even fewer surveys report SIA coverage along with information on prior measles vaccination status [35]. Other household surveys, such as Demographic and Health Surveys, try to capture specific information on measles vaccination [36], but the ability to compare SIA dose receipt among zero-dose or previously vaccinated children is constrained by the low proportion of children with documentation of routine vaccination and potential misclassification of routine and/or SIA vaccination when relying on maternal recall [36]. For each measles vaccination campaign, however, the size of zero-dose population reached by SIAs varies, depending on local routine coverage and SIA implementation approach [35].

Additionally, SIA coverage reported on the WHO-UNICEF Joint Reporting Form may be overestimated [35, 37, 38], possibly due to vaccinating non-target populations or not capturing the unreached populations. Future use of post-SIA surveys that include the vaccination history of recipients will improve our understanding of whether individual SIAs reach zero-dose children and the actual SIA coverage in the community. For MCV2 impact estimates based on actual (historical) coverage, given the purposeful selection of high burden countries with low coverage, this analysis is likely to underestimate the potential future role of MCV2 in controlling measles.

While DynaMICE is a dynamic transmission model that captures the indirect (herd) effect of vaccination, it is does not capture international case importation. Also, due to model limitation in simulating measles outbreaks in subnational areas, differentiation between outbreak response SIAs and preventative SIAs was not explored in this analysis. The potential impact of subnational variation in key determinants of measles transmission such as birth rates, routine and SIA vaccination coverage, and migration was also not assessed. Finally, NNV is not applicable for comparison between strategies when an alternative scenario does not avert additional cases, regardless of the number of doses administered. Our results show NNVs vary across countries, but there is no established threshold to determine whether efficiency of an immunisation programme is acceptable [39]. Further data, such as costs for vaccine procurement and delivery, will be useful in understanding cost-effectiveness of immunisation programmes.

The resources required for SIAs, including economic and human resources as well as logistical challenges, can be major deterrents to their implementation. Despite several unknowns regarding interpretation of estimated NNVs, our results show that routine MCV2 is not always more efficient than SIAs. Furthermore, the current trend towards including multiple interventions in a single SIA or integrating many of the components of SIA planning across different interventions may increase efficiency, although it will be important to monitor the effectiveness of “integrated campaigns” [40, 41].

We critically assessed the incremental impact of different measles vaccination strategies to inform future decisions about vaccination planning and policies. Understanding the relative impact and efficiency of the first routine dose, second routine dose, and SIAs of MCV will assist stakeholders in assessing the value of measles vaccination programmes, and further identify improved pathways towards measles elimination.

## Data Availability

Publicly available datasets were used for country models in this study, including measles vaccine coverage data from the World Health Organization (routine immunisation: https://immunizationdata.who.int/pages/coverage/mcv.html; supplementary immunisation activities: https://www.who.int/entity/immunization/monitoring_surveillance/data/Summary_Measles_SIAs.xls) and population statistics from the United Nations World Prospect Project 2019 (https://population.un.org/wpp/). Computer codes for simulating measles burden and vaccination strategies based on the DynaMICE model are available in https://github.com/hfu915/dynamice_ph.

## Authors’ contributions

MA, HF, KA, and MJ conceptualised the study. MA and HF curated the data, conducted the modelling analysis, prepared visualisations of results, and wrote the original draft. All authors contributed to reviewing results and editing of the manuscript and have approved the final version. The authors alone are responsible for the views expressed in this article and they do not necessarily represent the decisions, policy, or views of their affiliated organisations.

## Acknowledgements

This study was funded by the Bill & Melinda Gates Foundation (OPP1157270) and Gavi, the Vaccine Alliance.

This work was carried out as part of the Vaccine Impact Modelling Consortium (www.vaccineimpact.org), but the views expressed are those of the authors and not necessarily those of the Consortium or its funders. The funders were given the opportunity to review this paper prior to publication, but the final decision on the content of the publication was taken by the authors.

This work was supported, in whole or in part, by the Bill & Melinda Gates Foundation, via the Vaccine Impact Modelling Consortium [Grant Number INV-009125]. Under the grant conditions of the Foundation, a Creative Commons Attribution 4.0 Generic License has already been assigned to the Author Accepted Manuscript version that might arise from this submission.

