## Appendix for "Health impact of routine measles vaccination and supplementary immunisation activities in 14 high burden countries: a DynaMICE modelling study"

**Table S1. Number of cases across modelled countries 2010-2019 and vaccination schedules**

| Country | Reported number of cases (WHO/UNICEF) | Estimated number of cases (IHME) | Nationally recommended age for MCV1 | Nationally recommended age for MCV2 | Year of MCV2 introduction |
|---|---|---|---|---|---|
| India | 192566 | 112263832 | 9–12 months | 16–24 months | 2011 |
| Nigeria | 169394 | 15381229 | 9 months | 15 months | 2019 |
| Indonesia | 115974 | 15373892 | 9 months | 18 months | 2003 |
| Ethiopia | 59824 | 10108569 | 9 months | 15 months | 2019 |
| China | 213832 | 6171658 | 8 months | 18 months | 2005 |
| Philippines | 149325 | 6050672 | 9 months | 12–15 months | 2009 |
| Uganda | 20029 | 4555096 | 9 months | - | - |
| DRC | 791259 | 3976920 | 9 months | - | - |
| Pakistan | 74209 | 3271893 | 9 months | 15 months | 2009 |
| Angola | 30564 | 3246826 | 9 months | 15 months | 2015 |
| Madagascar | 234682 | 2966145 | 9 months | 15–18 months | 2020 |
| Ukraine | 129608 | 154625 | 12 months | 6 years | 2000 |
| Malawi | 118775 | 852873 | 9 months | 15 months | 2015 |
| Somalia | 80769 | 2440208 | 9 months | - | - |

**Table S1.** For countries without available data on the recommended MCV2 schedules, we assumed the age at vaccination to be 15–18 months old [8]. In China and Philippines, where multiple types of vaccines are in use, we adopted the vaccination schedules from the two doses of MMR (measles-mumps-rubella). Year of MCV2 introduction was assumed to be the first year a country has available coverage data in the WUENIC database. (-) indicates a country has not yet introduced MCV2 and therefore, does not have a recommended age for MCV2 vaccination. IHME: Institute for Health Metrics and Evaluation. MCV1: the first routine dose of measles-containing vaccine. MCV2: the second routine dose of measles-containing vaccine. WUENIC: WHO and UNICEF Estimates of National Immunization Coverage.

**Table S2. Averted deaths (thousands) across different vaccination strategies from 2000–2020**

| Country by MCV2 introduction year | Comparator: no vaccination | | Comparator: MCV1 alone | | |
|---|---|---|---|---|---|
| | MCV1 + MCV2 + SIAs | MCV1 alone | MCV1 + SIAs | MCV1 + MCV2 | MCV1 + MCV2 + SIAs |
| **MCV2 < 2017** | | | | | |
| India | 2393 | 2223 | 122 | 84 | 170 |
| Indonesia | 372 | 308 | 50 | 30 | 64 |
| China | 1546 | 1377 | 94 | 149 | 169 |
| Philippines | 199 | 160 | 35 | 9.82 | 39 |
| Pakistan | 1371 | 989 | 354 | 94 | 382 |
| Angola | 532 | 262 | 267 | 9.93 | 270 |
| Ukraine | 56 | 50 | 0.22 | 5.84 | 6.03 |
| Malawi | 333 | 264 | 69 | 5.69 | 69 |
| **MCV2 > 2017** | | | | | |
| Nigeria | 2812 | 1475 | 1337 | 5.19 | 1337 |
| Ethiopia | 1689 | 858 | 830 | 9.45 | 831 |
| Madagascar | 193 | 135 | 57 | 0.21 | 57 |
| **No MCV2** | | | | | |
| Uganda | 797 | 546 | 251 | 0 | 251 |
| DRC | 1328 | 841 | 488 | 0 | 488 |
| Somalia | 229 | 110 | 119 | 0 | 119 |
| **Total** | 13850 | 9598 | 4073 | 404 | 4251 |

**Table S2.** This table presents the total deaths averted in the 5 pairs of vaccination delivery strategies for comparison. Sums of the averted deaths in the 14 countries are shown in the last row of the table. DRC: Democratic Republic of the Congo. MCV1: the first routine dose of measles-containing vaccine. MCV2: the second routine dose of measles-containing vaccine. SIA: supplementary immunisation activity

**Table S3. Averted DALYs (thousands) across different vaccination strategies from 2000–2020**

| Country by MCV2 introduction year | Comparator: no vaccination | | Comparator: MCV1 alone | | |
|---|---|---|---|---|---|
| | MCV1 + MCV2 + SIAs | MCV1 alone | MCV1 + SIAs | MCV1 + MCV2 | MCV1 + MCV2 + SIAs |
| **MCV2 < 2017** | | | | | |
| India | 162378 | 150606 | 8427 | 5883 | 11772 |
| Indonesia | 26019 | 21582 | 3479 | 2144 | 4437 |
| China | 115911 | 103400 | 6943 | 11081 | 12511 |
| Philippines | 14066 | 11343 | 2447 | 701 | 2723 |
| Pakistan | 92423 | 66434 | 24065 | 6476 | 25989 |
| Angola | 30978 | 15144 | 15656 | 625 | 15834 |
| Ukraine | 3836 | 3435 | 15 | 387 | 400 |
| Malawi | 19019 | 15074 | 3939 | 371 | 3946 |
| **MCV2 > 2017** | | | | | |
| Nigeria | 150748 | 78181 | 72567 | 294 | 72567 |
| Ethiopia | 105623 | 54084 | 51513 | 643 | 51539 |
| Madagascar | 12442 | 8704 | 3730 | 15 | 3738 |
| **No MCV2** | | | | | |
| Uganda | 46851 | 32235 | 14616 | 0 | 14616 |
| DRC | 78817 | 49891 | 28925 | 0 | 28925 |
| Somalia | 13080 | 6205 | 6875 | 0 | 6875 |
| **Total** | 872191 | 616320 | 243197 | 28619 | 255872 |

**Table S3.** This table presents the total DALYs averted in the 5 pairs of vaccination delivery strategies for comparison. Sums of the averted DALYs in the 14 countries are shown in the last row of the table. DALY: disability-adjusted life year. DRC: Democratic Republic of the Congo. MCV1: the first routine dose of measles-containing vaccine. MCV2: the second routine dose of measles-containing vaccine. SIA: supplementary immunisation activity

**Table S4. Percentage of years over 2000–2020 showing a smaller number of susceptible children than the birth cohort size**

| Country | No vaccination | MCV1 | MCV1+ MCV2 | MCV1+SIAs | MCV1+MCV2+SIAs |
|---|---|---|---|---|---|
| India | 0% | 0% | 9.5% | 0% | 14% |
| Nigeria | 0% | 0% | 0% | 29% | 29% |
| Indonesia | 0% | 0% | 0% | 0% | 19% |
| Ethiopia | 0% | 0% | 0% | 14% | 14% |
| China | 0% | 0% | 67% | 24% | 67% |
| Philippines | 0% | 0% | 0% | 24% | 38% |
| Uganda | 0% | 0% | 0% | 38% | 38% |
| DRC | 0% | 0% | 0% | 0% | 0% |
| Pakistan | 0% | 0% | 0% | 14% | 33% |
| Angola | 0% | 0% | 0% | 24% | 24% |
| Madagascar | 0% | 0% | 0% | 29% | 29% |
| Ukraine | 0% | 0% | 0% | 0% | 0% |
| Malawi | 0% | 0% | 4.8% | 43% | 52% |
| Somalia | 0% | 0% | 0% | 9.5% | 9.5% |
| Median ($25^{th}$—$75^{th}$ percentiles) | 0% (0%—0%) | 0% (0%—0%) | 0% (0%—0%) | 18% (3%—26%) | 26% (14%—37%) |

**Table S4.** This table presents the percentage of years over the analysis period that had an outbreak potential among different vaccination strategies. The outbreak potential in a year is indicated by a larger size of susceptible population under 5 years old compared to the size of birth cohort in each country. The median and $25^{th}$ and $75^{th}$ percentiles of percentage among 14 countries are shown in the bottom row. DRC: Democratic Republic of the Congo. MCV1: the first routine dose of measles-containing vaccine. MCV2: the second routine dose of measles-containing vaccine. SIA: supplementary immunisation activity.

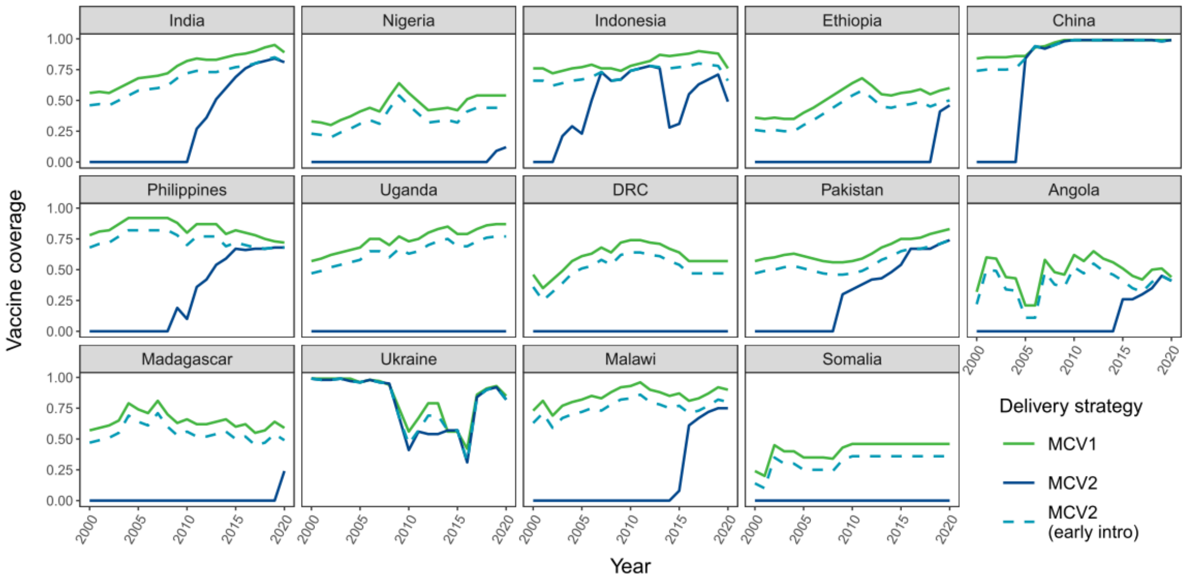


**Figure S1. Immunisation coverage for early introduction of MCV2 over 2000–2020.**
In the sensitivity analysis, all countries were assumed to introduce MCV2 early in 2000, with each year's coverage 10% lower than MCV1 coverage, or the same as the country's actual MCV2 coverage in that year, whichever was larger. DRC: Democratic Republic of the Congo. MCV1: the first routine dose of measles-containing vaccine. MCV2: the second routine dose of measles-containing vaccine.

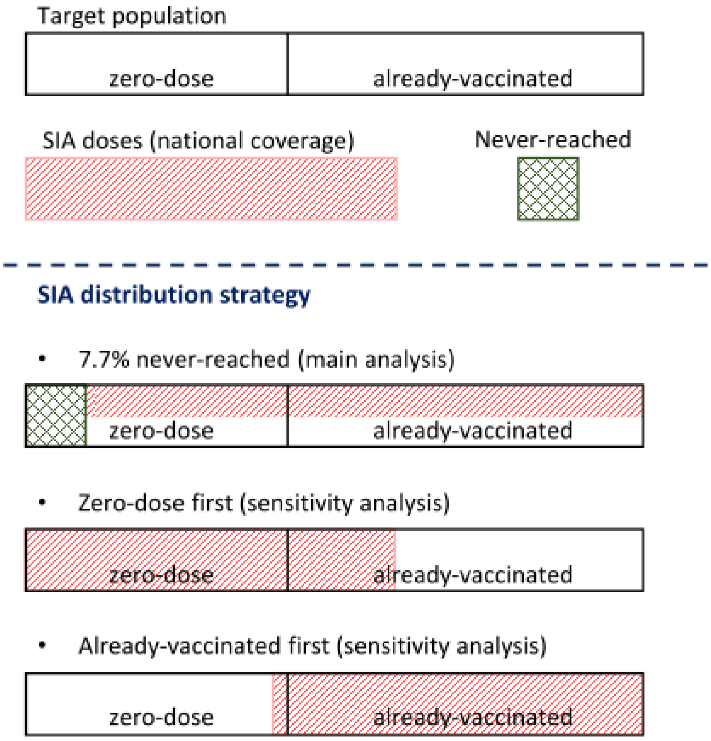


**Figure S2. Distribution strategies of SIAs included in the analysis.**
In the main analysis, 7.7% of target population are assumed to never receive MCV doses and SIAs doses are given randomly to the rest of population. In addition, the distribution strategies to direct SIA doses first to MCV zero-dose and already-vaccinated children are assessed respectively in the sensitivity analysis. MCV: measles-containing vaccine. SIA: supplementary immunisation activity.

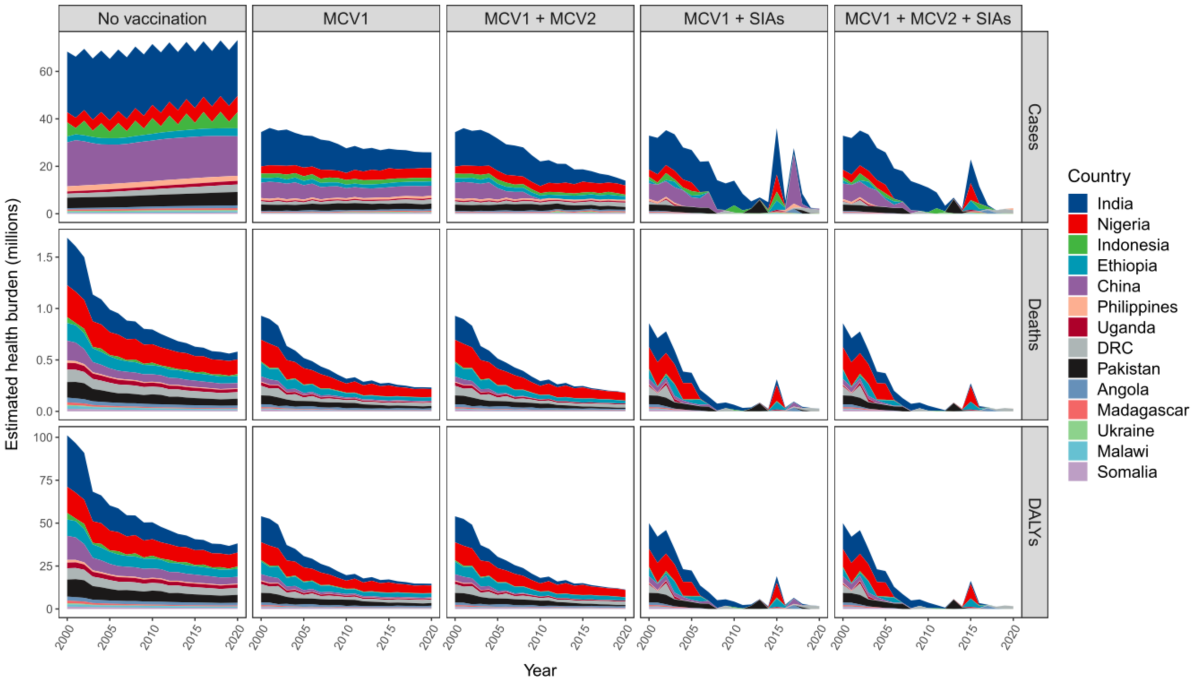


**Figure S3. Annual number of measles cases (top row), deaths (middle row), and DALYs (bottom row) across different vaccination delivery strategies over 2000–2020.**
Country measles burden is present in different colours and stacked over time. The measles burden decreases with adding vaccination delivery strategies. DALY: disability-adjusted life years. DRC: Democratic Republic of the Congo. MCV1: the first routine dose of measles-containing vaccine. MCV2: the second routine dose of measles-containing vaccine. SIA: supplementary immunisation activity.

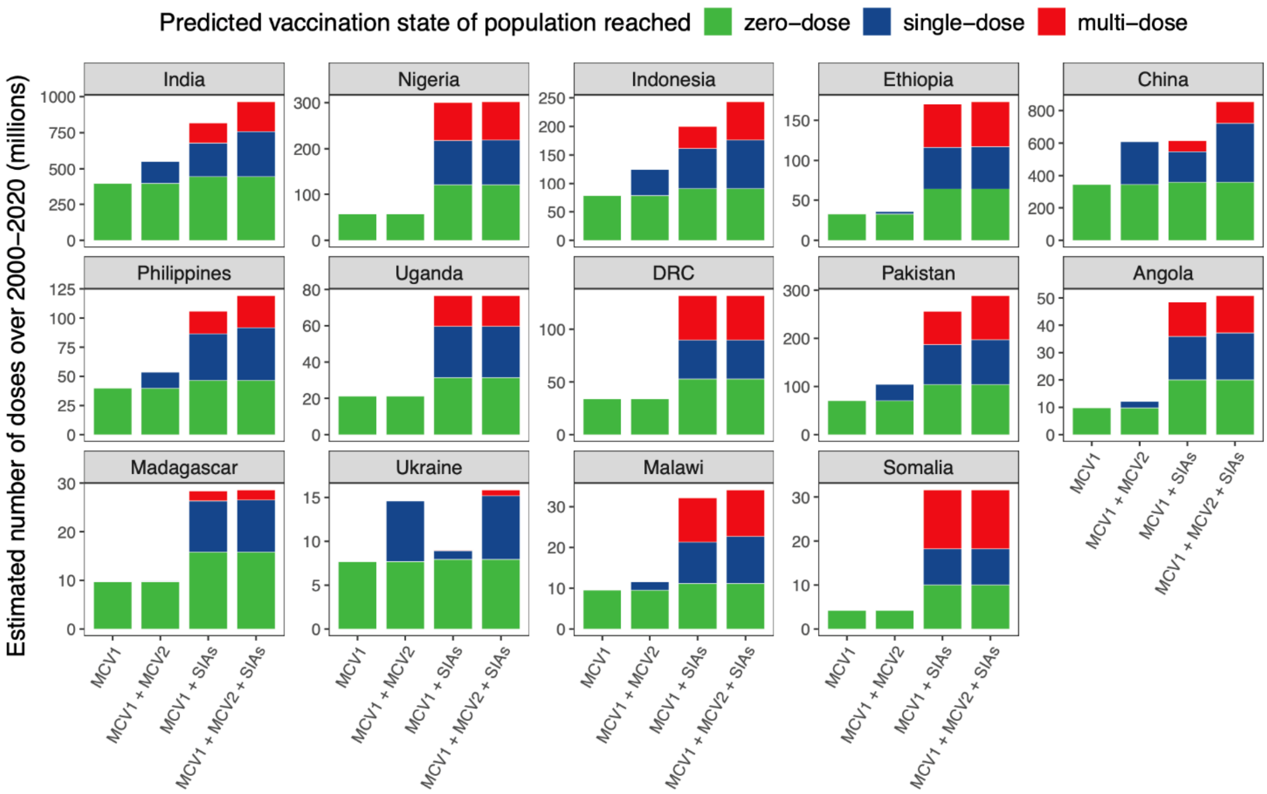


**Figure S4. Doses reaching zero-dose, single-dose, and multiple-dose children across different vaccination delivery strategies**

Total vaccine doses administrated over 2000–2020 are aggregated by estimated vaccination state of the target children reached. DRC: Democratic Republic of the Congo. MCV1: the first routine dose of measles-containing vaccine. MCV2: the second routine dose of measles-containing vaccine. SIA: supplementary immunisation activity.

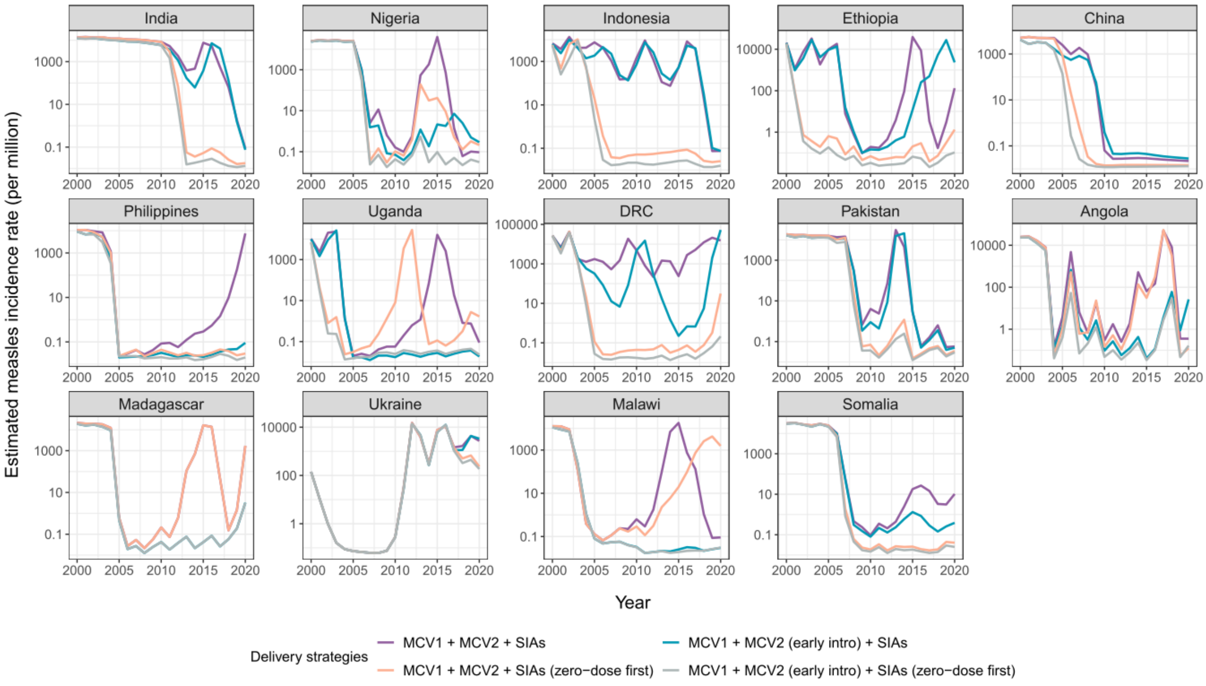


**Figure S5. Estimated measles incidence rate (per million) under alternative assumptions of delivering MCV2 and SIAs over 2000–2020.**

To estimate the impact of "optimal" vaccination impact (though not changing assumed MCV1 coverage or overall SIA coverage), we combined the alternative assumptions of early MCV2 introduction and SIA dose allocation prioritised for zero-dose populations. Incidence rates for different strategies are in different colours. Note that the y-axis is on the log scale. MCV2: the second routine dose of measles-containing vaccine. SIA: supplementary immunisation activity.
